# Personal and Social Determinants of Health Promoting Behavior of School going Adolescents in Pokhara, Nepal

**DOI:** 10.1101/2025.10.19.25338329

**Authors:** Ratna Shila Banstola, Usha Kiran Poudel, Kalpana Paudel Aryal, Indira Banstola, Sworup Bhandari, Sachiko Inoue

## Abstract

**Background:** Health-promoting lifestyle choices of adolescents are closely related to current and subsequent health status. Given the lack of understanding among Nepalese adolescents, the study is aimed to examine the health promoting behavior (HPB) and its personal and social determinants among school going adolescents in Pokhara, Nepal.

**Methods:** A cross-sectional study was conducted among 421 adolescents aged 11 to 18 years-old studying in class 6 to 10 of three randomly selected public schools of Pokhara Metropolitan. Structured self-administered questionnaire was used to collect data. Adolescent Health Promotion Scale – Short Form (AHP-SF), a 21 item Likert-scale was used which consists of six dimensions (nutrition, social support, health responsibility, life appreciation, physical activity, and stress management). Data analysis was done in SPSS 16 applying both descriptive and inferential statistics (linear regression analysis at <0.05 level of significance and 95% confidence interval).

**Results:** The mean score of HPB was 62.66±8.49 and 48.7% adolescents had inadequate level of HPB. Perceived school climate, play ground in school, social gathering, number of friends to play after school, parental concern/supervision, mother’s education, and family type determined the HPB. Perceived school climate strongly predicted all the dimensions of HPB: nutrition, social support, health responsibility, life appreciation, stress management and exercise. Individual predictors of each six dimensions of HPB are also identified.

**Conclusions:** Significant number of adolescents have inadequate HPB. Intervention to adolescents, parents, school and public can be planned focusing on the determinants of HPB identified by this study. The study has important policy and practical implication to enhance HPB and lead to healthful development of adolescent children.

## Introduction

Modern science largely reduced the risk of death from most infectious diseases, however, significant number of people are dying relatively young from non-communicable diseases (NCDs) [1]. Each year NCDs kill 41 million people accounting for 74% of all deaths globally, of those, 17 million people die from an NCD before the age of 70, with 86% of these premature deaths occurring in low- and middle-income countries (LMICs) [2–4]. There is double burden of morbidity and mortality because NCDs disproportionately affecting people in LMICS, where nearly three quarters of global NCD deaths occur [5,6]. Considering one of the LMICs in South-East-Asia region, in Nepal 71.1% of deaths were due to NCDs, of those deaths, 38.1% were due to behavioral risk factors [7]. Also inequities in health care access and services persists in LMICs such as Nepal [8]. Socioeconomic condition is closely linked with health of the people, for example, poor and vulnerable people have low access to health services and die sooner than those who can afford health care cost [5]. To see more critically, Nepal’s urban population has increased and reached 66.8%, as more people are migrating to cities led to mismanaged urbanization and change in family demography, indicating need of policies to develop urban infrastructure that can promote and maintain health of people [9, 10], however, still there is not much concern given to the healthy development of future citizens of Nepal, the children and adolescents’ need of school and community resources, and green spaces for their healthy development [11]. Although the countries are committed, there are many challenges to achieve the sustainable development goal of reducing one third of premature deaths from NCDs by 2030, because premature NCD mortality is increasing in LMICs, where the rates of diabetes and obesity are on the rise [1]. Behavioral risk factors are rising specifically the tobacco and harmful alcohol consumption, unhealthy diet and lack of physical activity [5]. The WHO extended global action plan for the prevention and control of NCDs from 2013–2020 to 2030, and called the nations for the development of an Implementation Roadmap 2023 to 2030 to accelerate progress on preventing and controlling NCD [5]. However, countries with low economy including Nepal are very far to reach these action plan and make the race on the roadmap for prevention and control. The path to success for NCDs is a long one, however, through the intervention based on evidences on the healthy behavior of generations to come may improve future quality of life and reduces health care cost [1].

In this regard, adolescence is a critical stage of development, while this age is thought a relatively healthy period of life, adolescents begin to make lifestyle choices and establish behaviors that affect both their current and future health as well as the behaviors may last the rest of their lives [6,12]. Therefore, helping adolescents establish healthy lifestyles and avoid developing health risk behaviors is crucial and should be started before these behaviors are firmly established [13,14]. A health-promoting lifestyle is a multi-dimensional pattern of self-initiated feelings and behaviors, such lifestyle includes eating a low-fat diet, maintaining nutrition, regular physical activities, maintaining a healthy body weight, avoiding smoking, developing good interpersonal relationships, reducing stress, which consequently helps to prevent many chronic diseases in the future [1,7,12]. Adolescents may not do alone, parents have an important role in helping their adolescents stay healthy, as parents can share a responsibility in teaching children about the links between food, health and environment [15]. Also there are other influences on the choices adolescents make, for instance, children and adolescents spend most of their time in school. Therefore, making schools and academic institutions an ideal place to foster lifelong healthy behaviors as well as other community level supports for example, family-peer-neighborhood environmental capital can also make the healthy choice the easy choice [16]. Relatedly, a study revealed urban neighborhood space environment can affect the duration of children’s physical activity [17]. Moreover, health maintenance and promotion are the fundamental prerequisites to the development. UNESCO and the World Health Organization launched the Global Standards for Health-promoting Schools on 22 june, 2021, a resource package for schools to improve the health and well-being of 1.9 billion school-aged children and adolescents [14]. However, information on how the family, neighborhood built structures, school climate and increasing sociodemographic transition in the urban areas of LMICs influences the HPB of school children and adolescents in terms of their nutrition, exercise, support seeking and relation, stress management, Life appreciation and health responsibility are lack.

Although few studies among other than Nepalese adolescents reported about HPB [18–21], there is gap of knowledge on factors that can have huge influence on establishing healthy lifestyle habits such as, self-efficacy a personal factor, and school and community related factors: school climate, neighborhood built environment (eg: playground, parks etc), parents’ role, together with changing family demographics in urban areas of LMICs. For example, a study among Turkish adolescents determined the inverse relationship between game addiction and HPB [22], and another reported positive relationship of self-efficacy levels and health literacy levels with health behaviors [23]. A study among German adolescents examined the impact of perceived school-climate and academic well-being on young people’s self-rated health [24]. Study from Belgium revealed perceived physical school environment were significant predictors of school-related physical activity (summation of in-school physical activity and active transport to school) [25]. Therefore, to add these limited understanding from developed context and none from Nepal on how is the influence of self-efficacy, school climate, parents’ support and built environment on children and adolescents’ HPB in LMICs needed to be explored. To fill this gap, we aimed to assess the personal factors (demographics and self-efficacy) and social factors (perception of school climate, family and neighborhood context, built environment, parents’ concern) and HPB among school going adolescents of Pokhara, Nepal.

## Methods

### Research design and participants

A cross-sectional study was conducted in three public schools of Pokhara metropolitan, Nepal. Pokhara is the second largest city in Nepal by area with total population of 518,000, and approximately 22% of the population is adolescents. There are more than 300 schools in Pokhara. The study population was adolescent students studying in three randomly selected public schools of Pokhara. Inclusion criteria was adolescent students studying in classes six to ten from allocated classes by school authority, whose parents gave written consent, who were present at the day of data collection, and were willing to participate in the study. Sample Size was calculated taking the prevalence of HPB of school going adolescents in India: 53% [18], using Cochran’s formula (n = z^2^pq/d^2^) where allowable error was kept at 5%. So that the sample size required was 383. The questionnaire was distributed to 432 students. Removing 11 questionnaire with incomplete responses, the final sample size for this study was 421.

### Measures

Structured self-administered questionnaire was used to collect the data. Questionnaire included socio-demographic information such as, age, sex, religion, ethnicity; family type, number of family members, education and occupation of parents, and currently living with.

#### School climate

A four-point likert scale from strongly agree to strongly disagree with 18 statements was used [26–29]. The items included were: school connectedness (I like this school, I feel like I am part of this school, I am happy to be at this school, I feel like I belong, Most days I look forward to going to school); peer social support (I get along with other students at school, Students in our school get along well, At this school, students talk about the importance of understanding their own feelings and the feelings of others, Students at my school treat each other with respect, Students in my school are welcoming to new students); Teacher support (It is easy to talk with teachers at this school, Teachers are available when I need to talk with them, My teachers care about me, Teachers understand my problems, My teachers make me feel good about myself, Teachers treat me with respect, Teachers treat all students fairly).

#### self-efficacy

Self-efficacy scale with 10 statements in four-point scale (not at all true to exactly true) [30–32]. The included were: I can always manage to solve difficult problems if I try hard enough, If someone opposes me, I can find the means/ways to get what I want, It is easy for me to stick to my aims and accomplish my goals, I am confident that I could deal efficiently with unexpected events, I know how to handle unforeseen situations, I can solve most problems if I invest the necessary effort, I can remain calm when facing difficulties because I can rely on my coping abilities, When I am confronted with a problem, I can usually find several solutions, If I am in trouble, I can usually think of a solution, I can usually handle whatever comes my way.

#### Parental concern/supervision

Parental concern/supervision was measured with 11 statements on parents/family concern to health habits in three point scale (Often to Never), which included items such as, parents don’t allow junk food for me, I don’t get pocket money to buy junk foods, I bring home made foods for tiffin, my parents and family don’t prefer fast foods, family members take meals together at home, my family grow vegetables in own kitchen garden, parents tell me for regular exercise, my parents do regular physical exercise/activities, my parents like me to be physically active [33–36].

#### Built environment and social capital

Family and neighborhood environment characteristics, social relationship, friendship and play in neighborhood were included. For example, Is there parks and play ground near to your home/neigborhood?; Does your school have enough place to play for students? Do you have good relation with your friends?

The reliability alpha value of parental concern, school climate and self-efficacy was 0.6, 0.86 and 0.93 respectively.

#### Adolescents’ health promoting behavior

Measured with Adolescent Health Promotion Scale – Short Form (AHP-SF) [37]. The AHP-SF is a 21 item Likert-type self-report instrument, the questionnaire is consisted of six dimensions (nutrition, social support, health responsibility, life appreciation, physical activity, and stress management) to measure health promoting lifestyles in adolescents. The psychometric properties of the AHP-SF, including item analysis, factor analysis, and reliability measures, were assessed in previous studies [38]. Based on pretesting of the instrument one item (Make an effort to feel interesting and challenged every day) was removed in this study. Cronbach alpha of the final data was 0.86. Factor analysis revealed that the three items in nutrition, four items in social support and relation, four items in health responsibility, three items in life appreciation, three items in stress management and three items in exercise dimensions were loaded ≥.4 to .86.

### Data collection procedure

Data was collected using structured self-administered questionnaire in the respective classrooms during their study hours as the time and venue allocated by the school. The researchers (RSB, IB and SB) themselves collected the data. The researchers explained the questionnaire, provided the guidelines and were present to clarify the queries of the adolescent students during the process. It took about 30 minutes to complete the questionnaire by the adolescent students.

### Ethical considerations

Ethical approval was obtained from the Institutional Review Committee of Tribhuvan University, Institute of Medicine, Maharajgunj, Kathmandu (493 (6-11) E2 079/080, April 21, 2023). Formal permission for data collection was obtained from respective schools. Informed written consent from parents and assent from adolescents was obtained prior to data collection. The researchers (RSB and IB) explained the purpose of the study and then assent from adolescents was obtained. Two days prior to data collection, the researcher visited the school and the assigned classes. An information sheet that included the details about the study title, purpose and procedure, and a consent form was sent to the parents with the adolescents which was facilitated by the school teachers. The students whose parents provided the written consent and those adolescents provided their assent were then included in the study. Anonymity was maintained by telling them to not to write their name or any identification numbers in the questionnaire. The participants were given full authority to withdraw their participation at any time during the investigation without providing a reason. Precaution was taken throughout the study in every step to safeguard the right and welfare of all respondents. Obtained data was used for research purpose only.

### Data Analysis

Data was analyzed in SPSS version 16 using descriptive statistics (frequency, percentage, mean and standard deviation) and inferential statistics i.e., simple and multiple linear regression analysis was done at <0.05 level of significance and 95% confidence interval.

## Results

Table 1 shows the demographic characteristics of the 421 adolescents. The mean age of the participant was 14.70 with standard deviation of 1.25, 52.7% were females, majority of their family follow Hindu religion (87.4%) and 34.4% were belonged to Mongolian ethnic group. Adolescents belonged to joint family were 62.2%, and 87.2% were living with their biological parents. About parents’ education, 40.4% of the fathers had completed secondary level and still 12.6% were illiterate, while 35.4% mothers had secondary level education and 29.1% were illiterate. Regarding the occupation, 25.2% fathers were involved in business or self-employment, and 43.9% of mothers were homemaker, and 69.6% family’s monthly income was just sufficient for livelihood.

**Table 1.** Demographic Characteristics of the Adolescent Children (n=421).

| Characteristics | Frequency | Percent |
| --- | --- | --- |
| **Age** |  |  |
| 11-14 years | 189 | 42.5 |
| 15-18 years | 242 | 57.5 |
| Mean ± SD = 14.70 ± 1.25 | | |
| **Sex** |  |  |
| Male | 201 | 47.7 |
| Female | 220 | 52.3 |
| **Religion** |  |  |
| Hindu | 368 | 87.4 |
| Buddha | 20 | 4.8 |
| Islam | 23 | 5.5 |
| Christian | 9 | 2.1 |
| Others | 1 | 0.2 |
| **Ethnicity** |  |  |
| Brahmin/Chhetri | 132 | 31.4 |
| Mongolian | 145 | 34.4 |
| Dalit | 105 | 24.9 |
| Others | 39 | 9.3 |
| **Family Type** |  |  |
| Single | 262 | 62.2 |
| Joint | 159 | 37.8 |
| **Living with** |  |  |
| Parents | 367 | 87.2 |
| With relatives/others | 54 | 12.8 |
| **Father's Education** |  |  |
| Illiterate | 53 | 12.6 |
| Basic | 109 | 25.9 |
| Secondary | 170 | 40.4 |
| University Level | 43 | 10.2 |
| Do not Know | 46 | 10.9 |
| **Mother's Education** |  |  |
| Illiterate | 92 | 21.9 |
| Basic | 115 | 27.3 |
| Secondary | 149 | 35.4 |
| University Level | 36 | 8.6 |
| Do not Know | 29 | 6.9 |
| **Father's Occupation** |  |  |
| Works at home (not employed) | 48 | 11.4 |
| Business/self-employed | 106 | 25.2 |
| Wage earner | 95 | 22.6 |
| Service/job | 30 | 7.1 |
| Agriculture | 25 | 5.9 |
| Others (eg: foreign) | 99 | 23.5 |
| Do not know | 18 | 4.3 |
| **Mother's Occupation** |  |  |
| Works at home/Home maker (not employed) | 185 | 43.9 |
| Business/self-employed | 63 | 15.0 |
| Wage earner | 53 | 12.6 |
| Agriculture | 44 | 10.5 |
| Service/job | 33 | 7.8 |
| Others (eg: foreign) | 33 | 7.8 |
| Do not know | 10 | 2.4 |
| **Economic Status** |  |  |
| Not sufficient | 46 | 10.9 |
| Just sufficient | 293 | 69.6 |
| Surplus | 78 | 18.5 |
| Do not know | 4 | 1.0 |

Almost 32% of the adolescent children did not have friends at their neighborhood. Most of them 95.7% reported that they have good relation with their friends, 41.6% reported they do not involve in social gathering and 27.6% do not meet/visit relatives’ house accompanying with their family. Similarly, 26.4% do not have good relation with their neighbors. Regarding play at school, 71.5% sometimes play but 12.6% never play at school. Similarly, at home and neighborhood, 59.4% want to play but only 37.5% play outdoor games after school in their neighborhood. Different reasons were provided for not playing afterschool i.e., 39.1% said they do not have time to play, 15.2% said no play area/ground/park near to their home. Even those who play afterschool said that they play at the road side and there is no large space or ground in their neighborhood and 81% wish for the open area or park in the neighborhood where they can play outdoor safely. (Table 2)

**Table 2.** Built environment, Social Relations and Use of Electronic Devices related Information of the Adolescent Children (n=421).

| Characteristics | Frequency | Percent |
| --- | --- | --- |
| Number of friends after school |  |  |
| No friend at neighborhood | 133 | 31.59 |
| 1-5 | 140 | 33.25 |
| 6 or more | 148 | 35.15 |
| Good relation with friends |  |  |
| Yes | 403 | 95.7 |
| No | 18 | 4.3 |
| Involve in social gatherings (cultural and religious activity) |  |  |
| Yes | 246 | 58.4 |
| No | 175 | 41.6 |
| Often meet or go to visit relatives |  |  |
| Yes | 305 | 72.4 |
| No | 116 | 27.6 |
| Relation (neighbors come to your house and you go to theirs') |  |  |
| Yes | 310 | 73.6 |
| No | 111 | 26.4 |
| Use of electronic devices and internet |  |  |
| Do not use | 37 | 8.8 |
| <1 hour | 204 | 48.5 |
| 1 - 3 hours | 117 | 29.4 |
| >3 hours | 56 | 13.3 |
| Play at school |  |  |
| Often | 67 | 15.9 |
| Sometimes | 301 | 71.5 |
| Never | 53 | 12.6 |
| Play outdoor games afterschool (around home/neighborhood) |  |  |
| Yes | 158 | 37.5 |
| No | 263 | 62.5 |
| Reason for not playing after school (n = 263) |  |  |
| No play area at home and neighborhood | 40 | 15.2 |
| No time to play | 103 | 39.1 |
| No friends to play | 73 | 27.7 |
| I do not like to play outdoor | 65 | 24.7 |
| Not responded | 7 | 2.6 |
| Is there park and play ground in your neighborhood? |  |  |
| Yes | 198 | 47.0 |
| No | 223 | 53.0 |
| Do you want a playground/park in your neighborhood where you can play outdoor? |  |  |
| Yes | 341 | 81.0 |
| No | 80 | 19.0 |

Regarding the level of health promoting behavior, 51.3% adolescent children studying in public schools of Pokhara metropolitan has adequate health promoting behavior while 48.7% have inadequate level of health promoting behavior. (Table 3)

**Table 3.**
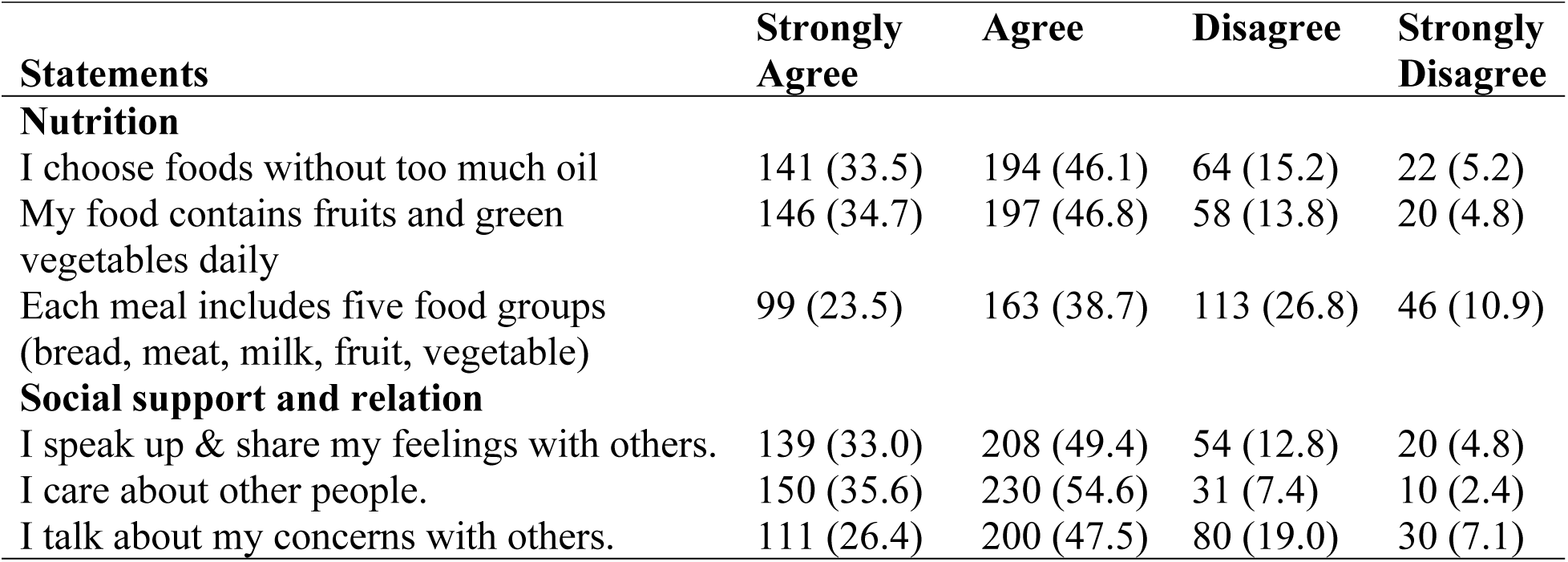
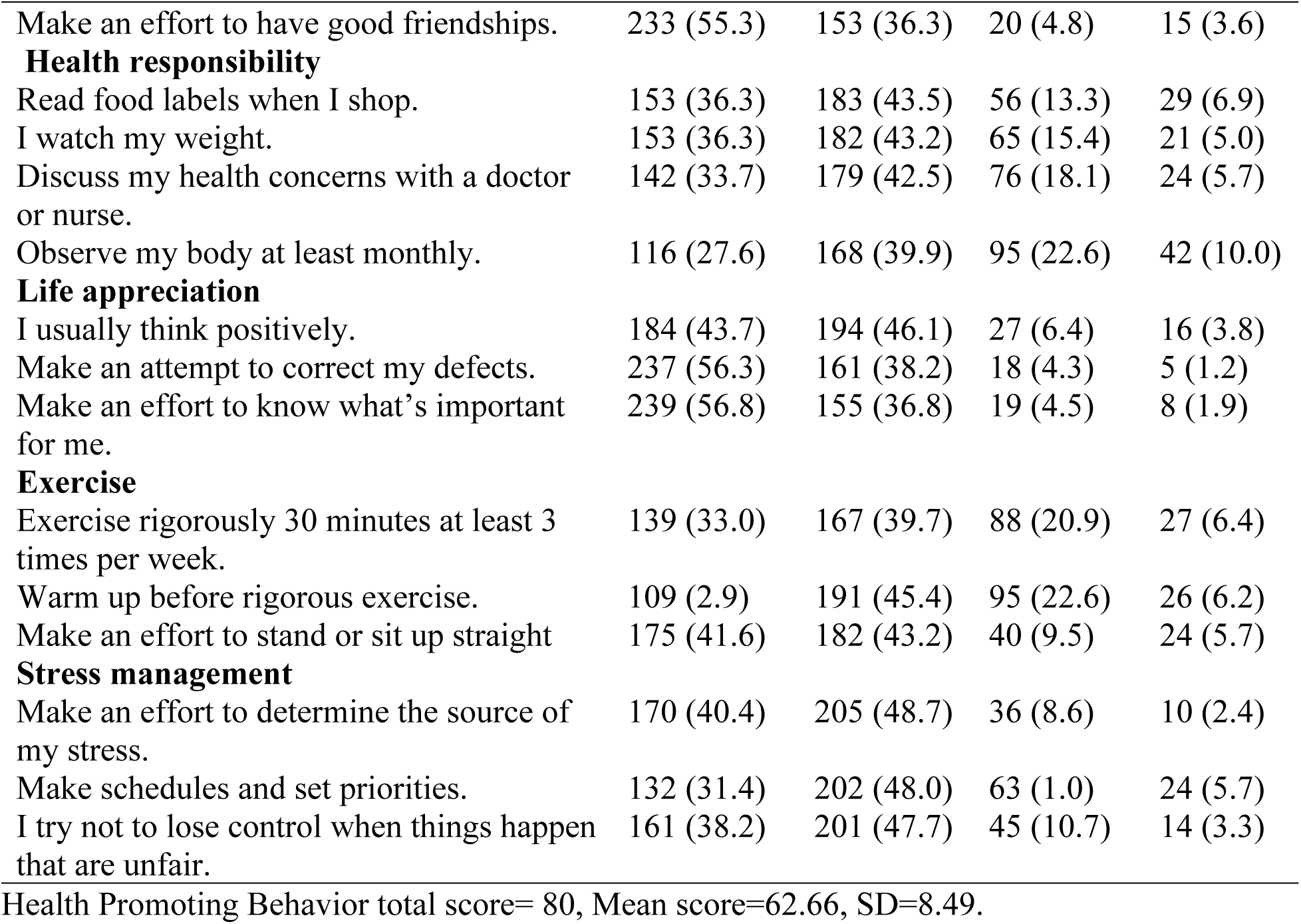
Responses on Health Promoting Behavior of the Adolescent Children (n=421).

The level of HPB was categorized based on mean score i.e., ≤mean as inadequate level of HPB and >mean as adequate HPB. Therefore, level of HPB: Adequate = 216 (51.3%); Inadequate = 205 (48.7%).

Multivariate linear regression revealed that family type (B = 1.967, *p* = .011, 95% CI = .450 to 3.483), mother’s education (B = -.064, *p* = .003, 95% CI = -.072 to .014), play ground in school (B = -4.123, *p* = .017, 95% CI = -7.500 to -.746), number of friends to play after school (B = -.219, *p* = .016, 95% CI = .040 to .397), social gathering at religious, cultural or community activities (B = -2.180, *p* = .006, 95% CI = -3.738 to -.622), parental concern (B = .329, *p* = .009, 95% CI = .084 to .575), and perceived school climate (B = .287, *p* = <.001 95% CI = .210 to .372) were the determinants of overall health promoting behavior of school going adolescents. (Table 4)

**Table 4.** Determinants of Health Promoting Behavior of the Adolescent Children (n=421).

| Variables | B | t | p | 95% CI |  |
| --- | --- | --- | --- | --- | --- |
|  |  |  |  | Lower | Upper |
| (Constant) | 45.139 | 5.707 | .000 | 29.589 | 60.690 |
| Age | -.513 | -1.678 | .094 | -1.114 | .088 |
| Sex | -.658 | -.832 | .406 | -2.212 | .897 |
| Religion | .629 | 1.304 | .193 | -.319 | 1.576 |
| Ethnicity | .319 | .813 | .417 | -.453 | 1.091 |
| Family type | 1.967 | 2.550 | **.011** | .450 | 3.483 |
| Father's education | .030 | 1.597 | .111 | -.007 | .067 |
| Mother's education | -.064 | -2.988 | **.003** | -.107 | -.022 |
| Father's occupation | -.029 | -1.343 | .180 | -.072 | .014 |
| Mother's occupation | .014 | .532 | .595 | -.037 | .064 |
| Economic status | .075 | 1.902 | .058 | -.002 | .152 |
| Living with | -1.184 | -1.286 | .199 | -2.995 | .626 |
| Play ground in School | -4.123 | -2.400 | **.017** | -7.500 | -.746 |
| Play at school | .238 | .328 | .743 | -1.188 | 1.664 |
| Play after school around home and neighborhood | 1.162 | 1.352 | .177 | -.527 | 2.850 |
| park and play ground in neighborhood | .738 | .961 | .337 | -.771 | 2.248 |
| Number of friends to play after school | .219 | 2.408 | **.016** | .040 | .397 |
| Relation with friends | .701 | .379 | .705 | -2.933 | 4.335 |
| Social gathering (cultural, religious, community) | -2.180 | -2.751 | **.006** | -3.738 | -.622 |
| Often meet/go to visit relatives | -1.429 | -1.614 | .107 | -3.169 | .311 |
| Relation with neighbor | .292 | .325 | .746 | -1.476 | 2.060 |
| Use of electronic devices | -.238 | -.721 | .471 | -.887 | .411 |
| Parental concern/supervision | .329 | 2.640 | **.009** | .084 | .575 |
| School climate | .287 | 6.592 | **.000** | .201 | .372 |
| Self-Efficacy | .082 | 1.296 | .196 | -.043 | .207 |

While we examined the six dimensions of HPB individually, linear regression stepwise model predicted that perceived school climate (B = .026, *p* = .005, 95% CI =.008 to .045), age (B = -.163, *p* = .019, 95% CI = -.299 to -.027), family type (B = .447, *p* = .013, 95% CI =.094 to .800), living with (B = -.565, *p* = .009, 95% CI = -.991 to -.139), play ground in school (B = -1.036, *p* = .010, 95% CI =-1.823 to -.248), parental concern/supervision (B = .066, p=.021, 95% CI =.010 to .123) and religion (B = .229, *p* = .043, 95% CI =.007 to .450) has significant effect on nutrition behavior of adolescents. Regarding the predictors of social support behavior, only perceived school climate showed significant positive effect (B = .072, *p* =.000, 95% CI =.052 to .092). The school climate (B = .065, *p* =.000, 95% CI = .041 to .089), social gathering at cultural, religious, community activities (B = -.702, *p* =.002, 95% CI = -1.151 to -.253), parental concern (B = .100, *p* =.006, 95% CI =.029 to.171), mother’s education (B = -.013, *p* =.005, 95% CI = -.021 to -.004), number of friends to play afterschool in neighborhood (B = .061, *p* =.012, 95% CI =.014 to .108), play ground in school (B = -1.167, *p* =.021, 95% CI = -2.161 to -.173), father’s occupation (b= -.012, *p* =.029, 95% CI = -.024 to -.001), family type (B = .482, *p* =.033, 95% CI =.040 to .925), often meet/go to visit relatives (B = -.520, *p* =.039, 95% CI = -1.015 to -.025) predicted the health responsibility behavior of adolescent children. Likewise, school climate (B = .039, *p* = <.001, 95% CI = 023 to .054) and self-efficacy (B = .028, *p* = <.001, 95% CI = .004 to .052) were the strong predictors of life appreciation behavior followed by social gathering at cultural, religious, community activities (B = -.345, *p* =.025, 95% CI = -.646 to -.043), and ethnicity (B = .165, *p* =.032, 95% CI =.014 to .316). Concerning stress management behavior, higher the perceived school climate has positive effect on stress management (B = .048, *p* = <.001, 95% CI = .031 to .064). Similarly, economic status (B= .019, *p* = .025, 95% CI = .002 to .035) and self-efficacy (B = .029, *p* = .025, 95% CI = .004 to .054) also showed positive effect, while the use of electronic devices negatively predicted the stress management (B = -.183, *p* = .009, 95% CI = -.319 to -.047). Regarding exercise behavior, perceived school climate showed the most positive effect (B = .055, *p* = <.001, 95% CI = .035 to .075). While age, sex and social gathering predicted negatively (B = -.153, *p* = .043, 95% CI = -.302 to -.005; B = -1.013, *p* = <.001, 95% CI = -1.385 to -.641; and B = -.554, *p* = .004, 95% CI = -.931 to -.177 respectively). Other predictors were mother’s education (B = -.013, *p* = .001, 95% CI = -.020 to -.005), parental concern/supervision (B = .083, *p* = .007, 95% CI =. 023 to .143), family type (B = .444, *p* = .021, 95% CI = .068 to .821), and economic status (B = .022, *p* = .024, 95% CI = .003 to .042). (Table 5)

**Table 5.** Predictors of Six Sub-scales of Health Promoting Behavior (n=421).

| Variables | B | p | 95% CI |  |
| --- | --- | --- | --- | --- |
|  |  |  | Lower | Upper |
| **Nutrition Behavior** |  |  |  |  |
| (Constant) | 8.925 | <.001 | 6.161 | 11.688 |
| School climate | .026 | .005 | .008 | .045 |
| Age | -.163 | .019 | -.299 | -.027 |
| Family type | .447 | .013 | .094 | .800 |
| Living with | -.565 | .009 | -.991 | -.139 |
| Play ground in School | -1.036 | .010 | -1.823 | -.248 |
| Parental concern/supervision | .066 | .021 | .010 | .123 |
| Religion | .229 | .043 | .007 | .450 |
| **Social Support** |  |  |  |  |
| (Constant) | 8.489 | <.001 | 7.297 | 9.681 |
| School climate | .072 | <.001 | .052 | .092 |
| **Health Responsibility** |  |  |  |  |
| (Constant) | 7.943 | <.001 | 5.207 | 10.679 |
| School climate | .065 | <.001 | .041 | .089 |
| Social gathering (cultural, religious, community) | -.702 | .002 | -1.151 | -.253 |
| Parental concern/supervision | .100 | .006 | .029 | .171 |
| Mother's education | -.013 | .005 | -.021 | -.004 |
| Number of friends to play afterschool in neighborhood | .061 | .012 | .014 | .108 |
| Play ground in School | -1.167 | .021 | -2.161 | -.173 |
| Father's occupation | -.012 | .029 | -.024 | -.001 |
| Family type | .482 | .033 | .040 | .925 |
| Often meet/go to visit relatives | -.520 | .039 | -1.015 | -.025 |
| **Life Appreciation** |  |  |  |  |
| (Constant) | 7.335 | <.001 | 6.063 | 8.607 |
| School climate | .039 | <.001 | .023 | .054 |
| Self-efficacy | .028 | .021 | .004 | .052 |
| Social gathering (cultural, religious, community) | -.345 | .025 | -.646 | -.043 |
| Ethnicity | .165 | .032 | .014 | .316 |
| **Stress Management** |  |  |  |  |
| (Constant) | 6.138 | <.001 | 4.982 | 7.295 |
| School climate | .048 | <.001 | .031 | .064 |
| Use of electronic devices | -.183 | .009 | -.319 | -.047 |
| Economic status | .019 | .025 | .002 | .035 |
| Self-efficacy | .029 | .025 | .004 | .054 |
| **Exercise** |  |  |  |  |
| (Constant) | 7.772 | <.001 | 4.705 | 10.838 |
| School climate | .055 | <.001 | .035 | .075 |
| Sex | -1.013 | <.001 | -1.385 | -.641 |
| Social gathering | -.554 | .004 | -.931 | -.177 |
| Mother education | -.013 | .001 | -.020 | -.005 |
| Parental concern/supervision | .083 | .007 | .023 | .143 |
| Family type | .444 | .021 | .068 | .821 |
| Economic status | .022 | .024 | .003 | .042 |
| Age | -.153 | .043 | -.302 | -.005 |

## Discussion

The study was aimed to identify personal and social determinants of health promoting behavior of adolescent students aged between 11 to 18 years old, studying in public schools of Pokhara, Nepal. We also analyzed the predictors of six dimensions of HPB: nutrition, social support, health responsibility, life appreciation, stress management and exercise behavior.

Study found that the mean score of HPB was 62.66±8.49, which is similar to the findings among Iranian adolescents [39]. Likewise, based on mean score when categorized the level of HPB, it was found that 51.3% adolescents had adequate, while 48.7% had inadequate level of HPB. This significant number of adolescents with inadequate level of HPB shows that there is urgent and important need of interventions to be focused at family, community and school level to improve adolescents’ HPB.

Various determinants of HPB was identified in this study, includes demographic factors (mother’s education, family type), school climate, social gathering at cultural or community activities, parental concern or supervision, number of friends to play at neighborhood, and play ground in school. Discussing the demographic factors, children living in joint family were more likely to score higher on HPB than their counterparts with its beta coefficient 1.967. However, a previous study reported no any significant association of demographic factors including family structure of the students [40]. Demographic factors are country and culture specific variables, therefore, we added that family type has effect on Nepalese adolescent’s HPB. Concerning mother’s education, a study in Iran also reported significant association of the adolescent health promotion scale with age, gender, father’s educational level, mother’s educational level and mother’s occupation [39], however, we did not find the association with father’s education and mother’s occupation. Again this difference can be attributed by difference in country context. In our study those with low level of mother’s education were less likely to HPB, which aligns with previous findings from Egypt and Turkey [21,41]. Therefore, we added that demographic factors related to parents are important determinants of HPB in adolescents. Though we found if mothers are educated, they can contribute to HPB of their children, we also observed that despite the study was conducted in the second largest city of Nepal, using probability random sampling 29.1% of mothers were illiterate. Therefore, to improve HPB of children the policy and planning should also be focused to support education of women. We claim that educated mother can contribute to healthful development of their children who are the future adults.

Regarding social determinants of HPB, we observed that school climate is an important factor. School climate (school connectedness, peer support and teacher support together) was the strong predictor of HPB, which aligns with previous findings in different countries [42,43]. In addition, this study revealed that social gathering at cultural or community activities by the adolescents had the coefficient of -2.18, i.e., those who do not involve in social gathering were tend to have low score on HPB. In this regard, the importance of social relations and engagement on health promotion has been indicated by Simons-Morton et al. in their book Behavior theory in health promotion practice and research [44]. We also identified parental concern or supervision determines HPB i.e., each unit increase in parental concern score caused 0.329-unit increment in HPB score of their children. Scholars reported that parents and family have the great influence on health promotion and behavior development of their children [45]. Similarly, a Korean study also indicated parenting behavior and relationship predicts HPB in children [46]. Therefore, parents and family are the primary role model for their children to learn health habits.

Furthermore, built environment such as, play ground or park are important infrastructure where the children can do physical activities and play with friends but we found that only 37.5% play outdoor games after school in their neighborhood. Different reasons reported for not playing afterschool were, no time to play, no play-ground/park near to their home and no friends to play with etc. Even those who play reported that they play at the road side and there is no green space or ground in their neighborhood. Importantly, 81% adolescents expressed their wish for open area or park in the neighborhood where they can play outdoor safely. In the context of rapid urbanization with lack of urban planning in LMICs including Nepal [47,48], we suggest it is very urgent issue to be addressed considering the developmental requirement of its children and adolescents. Physical activities and play are one of the important requirement for health promotion and healthy development of children, therefore, considering these findings the government, municipality and the civil society need to work together and develop plan and policies in contributing healthy development of future citizen of the nation.

Similarly, our study revealed number of friends at neighborhood to play after school negatively determined the HPB and past study also reported friends are important part of socialization, development and learning of health related behavior. Another study also revealed that peer relationship as interpersonal factor predicts HPB of children [46]. Social relations, communication, and participation in socio-cultural and religious activity are great platform for the children to be socialized and strong medium to learn health habits and life skills. Unfortunately, 41.6% of the adolescent children reported they do not involve in social gathering and 27.6% do not often visit their relatives’ house accompanying with their family. Similarly, 26.4% reported not having good relation with their neighbors. Therefore, the finding signifies the need for targeted community intervention that can foster social cohesion and social capital. The nurses and health care workers can advocate and plan the intervention targeting positive social capital development that consequently contribute to health promotion, positive development and socialization of children and families in urban communities of LMICs.

Likewise, the effect of play and playground at school has been identified by previous studies from different countries [50–52]. Although most of the public schools in Nepal have playground (95%), only 71.5% sometimes play at school and 12.6% never play. Moreover, we revealed that those who reported no playground at school had reduced HPB with its coefficient of -4.123. In this regard, it is important that besides academic activities, play is very crucial for overall development of children. Nepalese education system can focus this finding, and revise the school activities to enhance children’s HPB.

### Predictors of individual dimensions of health promoting behavior

Nutritional behavior of adolescents was predicted by religion, family type, living with, parental concern/supervision, perceived school climate, and play ground in school. Those belonged to Hindu religion compared to others, those living in joint family, whose parents give concern to health habits or supervise, and those with higher perceived school climate were more likely to have higher score on nutritional behavior. While, those currently living with other than their biological parents were less likely to nutrition behavior. In support it is previously reported that parents’ behavior, encouraging their children for healthy eating and parental monitoring influences good nutrition habit in their children [33,34]. Additionally, a study from Southern Ecuador reported that parental permissiveness increased the soft drinks and unhealthy food habits in adolescents [53]. The studies among the adolescents in Southern Ecuador and Portugal partially supported our findings of school climate has effect on nutrition habit of their students [53,54]. Similarly, the influence of religion and joint family system on food and nutrition is well discussed [55]. The Centers for Disease Control and Preventions have also emphasized the role of parents, family and school for healthy nutrition habit in children [12].

Concerning social support behavior, only perceived school climate showed significant positive effect. This finding partially to fully agree with previously reported findings from three studies in USA and one from Iran respectively [28,42,43,56].

Current study found that health responsibility behavior of adolescents was determined by school climate, social gathering at cultural-community activities, parental concern/supervision, mother’s education, number of friends to play afterschool in neighborhood, play ground in school, father’s occupation, family type, often meet/go to visit relatives. The number of friends and influence of friendship relationship on HPB of children was also identified in a Korean study [46]. A scoping review has clearly emphasized that family could be the modeling and influencing mechanism to adopt healthy lifestyles by its members [57]. Additionally, self-efficacy as the individual level mechanism influences HPB [57]. In lack of previous studies that revealed the relation between school climate, and social capital (community participation and relationship in neighborhood) and health responsibility behavior of adolescents, we identified that school climate and social relationship has strong influence on health responsibility of adolescents. Therefore, these findings can be focused in future studies and in planning interventions.

Likewise, concerning the life appreciation behavior of adolescent children, the school climate and self-efficacy were found the strong predictors followed by social gathering and ethnicity. In line with the findings on the role of self-efficacy, a study among South Korean adolescents found that higher self-control was influencing factors of HPB [20]. Similarly, another study also reported the influence of self-efficacy on HPB of Korean children [46].

We also found that perceived school climate is strong predictor i.e., school climate has positive effect on stress management, and the use of electronic devices has negatively predicted the stress management behavior. In partial support of findings related to perceived school climate previous study among US adolescent reported higher health behavior [43], and some other studies also partially support our findings of stress management [42,56]. In support of the findings related to use of electronic devices a past study also identified the association of spending more time watching TV and playing computer games with lower AHP-SF scores in adolescents [37].

Again we revealed that perceived school climate showed the most positive effect on exercise behavior, while age, sex and social gathering predicted negatively. Other predictors identified were mother’s education, parental concern/supervision, family type and economic status. In agreement with the findings related to school climate study among Georgian adolescents also reported that the odds of being physically active increased with a more positive report of supportive school environments, school connectedness, peer social support, school physical environments, cultural acceptance, school safety, and adult social support [28]. To support our findings on sex as predictor, WHO indicted that adolescent girls were less active than adolescent boys, with 84% versus 78% not meeting the WHO recommendation of 60 minutes of activity per day [58]. A South Korean study also found that having higher economic status was related to better HPBs among adolescents [20]. Hence, we revealed that school climate strongly influences all the dimensions of HPB of adolescents. While, parents have significant role in nutrition, health responsibility and exercise behavior. Likewise, social capital (friends, neighborhood, social participation, often vising relatives) determines health responsibility, life appreciation and exercise behavior. At individual level (personal) mechanism, self-efficacy influenced two of the HPB’s life appreciation and stress management. Hence, our finding suggested that social determinants such as family, parents, school climate, neighborhood and social capital in terms of social participation and relationship strongly influences most of the health promotion behaviors of adolescents.

### Strengths and Limitations

This study was based on the primary data collected with a probability sampling technique. To our knowledge it is the first study which examined the determinants of health promoting behavior including the predictors of six dimensions of HPB among adolescents in Nepal. The study has filled the gap by adding in extant literature, understanding on personal and social determinants of HPB among adolescents from an LMICs from the global south context. Despite these strengths, there are some limitations of the study. It was based on self-report measure, thus might cause the social desirability effect or information bias. As it was the cross-sectional study, it is limited to establish a causal relationship.

## Conclusion

It is concluded that nearly half of adolescent have inadequate HPB. The determinants of HPB of adolescent children were perceived school climate, social gathering, mother’s education, parental concern, family type, number of friends to play after school, and play ground in school.

Regarding the dimensions of HPB, perceived school climate was the strong predictor of all the dimensions of HPB (nutrition, social support, health responsibility, life appreciation, stress management and exercise behavior), whereas self-efficacy predicted life appreciation and stress management behavior. Among the structural capital, playground predicted nutrition and health responsibility. Among variables related to social relationship, social gathering predicted health responsibility, life appreciation and exercise. Family system determined nutrition, health responsibility and exercise behavior, and living with parent positively affected nutrition. Parents’ concern determined the nutrition, exercise and health responsibility. The number of friends and often visit relatives predicted the health responsibility behavior of adolescent children. The school and family level intervention can be planned to enhance HPB in adolescent children. Thus the findings can be utilized by parents, schools, health professionals including school health nurses and those who work in promotion of health of children and adolescents which can have long term effect on health status and reduction of Non-communicable disease burden in the future.

## Data Availability

All relevant data are within the manuscript.

## Acknowledgments

We appreciate the participating schools for granting permission to study, and would like to thank all the adolescent students who participated in this study and provided their invaluable information.

## Author Contributions

**RSB:** conceptualization; methodology; data collection and curation; data analysis and writing original draft. **UKP:** literature review; methodology; writing manuscript. **KPA:** methodology; manuscript review. **IB:** data collection; data curation, writing-original draft. **SB:** literature review; data collection; data entry and analysis; writing manuscript. **SI:** methodology; data analysis; writing-review and editing.

## Data availability statement

The data are available from the corresponding author upon reasonable request.

